# Oxidative DNA Damage Diminishes After Treatment of Unipolar and Bipolar Depressive Episodes: A Systematic Review and Meta-Analysis of Longitudinal Studies

**DOI:** 10.1101/2025.05.20.25327903

**Authors:** Buket Yeşiloğlu, Ceren Gör, Reyhan Nur Babalıoğlu, Şevin Hun Şenol, Mark Frye, Anders Jørgensen, Ayşegül Özerdem, Deniz Ceylan

## Abstract

**Introduction:** Unipolar (UD) and bipolar depression (BD) have been associated with increased oxidative DNA damage, as measured by 8-hydroxy-2′-deoxyguanosine (8-OHdG) levels. While existing meta-analyses rely on cross-sectional data, this study aims to systematically review and meta-analyze existing longitudinal studies on oxidative DNA damage changes after treating UD and BD.

**Methods:** A literature search for 8-OHdG with UD and BD studies was conducted in PubMed, Cochrane, Scopus, Web of Science, and Ovid-MEDLINE until 31/12/2024. Screening was performed via Covidence and statistical analyses were performed in SPSS 28.0 using Hedges’s g (95% CIs) under a random-effects model, assessing heterogeneity (I^2^≥50%), publication bias (Egger’s test, funnel plots), and subgroup differences. Additionally, meta-regression on sex (%female) was performed.

**Results:** Out of 1512 studies, 10 met eligibility criteria, including 525 individuals (UD=376, BD =149). Meta-analysis revealed a post-pharmacotherapy reduction in 8-OHdG levels (g=-0.61, [−1.20, −0.02], p=0.04) with high heterogeneity (I^2^=0.95, Q=207.45, df(Q)=17, p<0.001) but no publication bias. In remitted patients (n=117), 8-OHdG levels significantly decreased (g=-0.42, [−0.81, −0.03], p=0.03). Subgroup analyses showed a significant decrease in BD population (g=-0.39, [−0.73, −0.05], p=0.02) with relatively high homogeneity (I^2^=0.49, Q=9.853, df(Q)=5, p=0.080). Significant reductions were observed in studies of pharmacological treatments (g=-0.88, [−1.56, − 0.19], p=0.001) and studies using urine samples (g=-0.61, [−0.84, −0.39], p<0.001). Moreover, meta-regression showed female sex as a significant moderator in the UD population (β=0.099, [0.065, 0.133], p<0.001).

**Conclusion:** Our findings support 8-OHdG levels as a potential biomarker in BD for treatment response. Further longitudinal studies are needed to validate these findings and to evaluate treatment-related changes.

## Introduction

Bipolar disorder (BD) and major depressive disorder (MDD) are chronic and highly disabling psychiatric conditions characterized by recurring mood episodes [1]. Previous studies suggested associations between mood disorders and an increased risk of many medical conditions, including cardiovascular disease, diabetes, cancer, stroke, early aging, and premature mortality [2-4]. While the biological mechanisms underlying mood disorders and treatment outcomes remain not completely elucidated, frequent medical comorbidities prompted investigations on potential shared mechanisms, such as oxidative stress and related pathways [5-7].

Oxidative stress arises when the balance between the generation and elimination of the reactive oxygen species (ROS) – the highly reactive byproducts of cellular oxidative respiration – is disrupted, leading to surpassing antioxidant defenses and lipid, protein, and DNA damage. Oxidative stress is associated with various diseases, including mood disorders [8-10]. Substantial evidence on mood disorders highlights alterations in stable oxidative end-products, such as lipid peroxidation and DNA damage markers, observed across different specimens – including blood, urine, and brain tissue – through various analytical techniques [11-13].

Studies that investigated the effect of pharmacological interventions, particularly antidepressants on oxidative stress markers led to evidence suggesting modulation of oxidative stress through antioxidant mechanisms as a possible mechanism underlying the mood regulatory effect of these medications. Previous meta-analyses suggested that oxidative stress indicating marker – malondialdehyde (MDA) – level was decreased after antidepressant treatment, while antioxidants such as uric acid and zinc levels have demonstrated an increase in MDD. On the other hand, data indicates that medicated patients with BD had higher levels of oxidative DNA damage than unmedicated patients, suggesting that certain medications may contribute to increased oxidative stress [14].

Oxidative DNA damage, a consequence of oxidative stress, significantly contributes to impairments such as cell death and mutagenesis. Increased oxidative DNA damage has been observed in both peripheral and brain samples in mood disorders, primarily measured through markers like DNA strand breaks and the oxidative end-product 8-hydroxy-2’-deoxyguanosine (8-OHdG) [15, 16]. 8-OHdG is the most frequently used marker of DNA damage arises from the oxidation of the highly vulnerable DNA guanine base, which is more commonly affected than other bases, and is easily detectable using several techniques [17, 18]. Successive meta-analyses highlight changes in 8-OHdG levels in individuals with mood disorders and even in populations at risk for BD [19, 20, 14, 21, 22, 7]. According to previous meta-analyses, the oxidative DNA damage biomarker 8-OHdG is elevated in depression compared to healthy controls [20, 21], with one meta-analysis specifically reporting that 8-OHdG/8-oxo-dG levels are increased during depressive episodes but not in manic or euthymic states in BD, suggesting that DNA damage may serve as an episode-related rather than a trait marker [22].

Moreover, a large sample size study presented lower 8-OHdG levels in MDD patients who employed antidepressant therapy in contrast to non-users regardless of being remitted or in an acute episode, which suggests that antidepressants might have an antioxidant effect [23]. In addition to these cross-sectional findings, accumulating recent longitudinal follow-up data have shown a sustained change in oxidative stress biomarkers, including oxidative DNA damage, following several treatments, particularly treatment with antidepressants [24-28].

Given the established links between mood disorders and oxidative stress biomarkers, investigating the effects of treatment on oxidation-induced DNA damage is crucial. While evidence suggests an effect of antidepressants and mood stabilizers, as well as electroconvulsive therapy (ECT), on oxidative DNA damage, the scarcity of longitudinal studies prevents meta-analytic evaluations of the long-term impact of acute episodes or remission on 8-OHdG levels. Findings remain inconsistent, with one study reporting increased 8-OHdG concentrations after follow-up in depressive patients [29], while others show contrasting results [25, 26, 30, 27, 28]. These discrepancies raise the question of whether treatments can modulate the imbalance between ROS and antioxidants. To date, no meta-analyses have specifically examined the effects of treatment on 8-OHdG levels.

Therefore, we conducted a systematic review and a meta-analysis study comparing 8-OHdG levels pre-versus post-intervention in unipolar and bipolar depressive episodes. In addition, this study aimed to investigate the association between oxidative DNA damage and subgroups such as different diagnostic groups, treatment modalities, biological specimens, and measurement methods that were used for quantification of 8-OHdG levels. Also, data from remitted patients were analyzed and the moderator effect of sex on 8-OHdG changes was assessed via meta-regression.

## Methods

This systematic review and meta-analysis have been registered in PROSPERO (Registration ID: CRD42025630675) and conducted according to The Preferred Reporting Items for Systematic Reviews and Meta-analyses (PRISMA). A literature search was held with the assistance of a librarian from Koç University, using the electronic databases of PubMed, Cochrane, Scopus, Web of Science, and Ovid MEDLINE from inception to December 31, 2024. The search keywords were (depression OR depressive) AND (8-hydroxy-2′-deoxyguanosine OR 8-hydroxydeoxyguanosine OR 8-hydroxy-deoxyguanosine OR 8-hydroxy-2-deoxyguanosine OR 8-hydroxydeoxyguanine OR 8-hydroxyguanine OR 8-oxoguanine OR 8-oxo-7,8-dihydro-2-deoxyguanosine OR 8-oxo-7,8-dihydro-guanosine OR 8-OHdG OR 8-OH-dG OR 8-OHdG OR 8OHdG OR 8-oxoGuo OR 8-oxo-dG OR 8-oxo-G OR 8-oxoG OR 8-OHG OR 8OH2’dG OR OH8dG OR “DNA damage”).

All articles found through this search were uploaded to Covidence; the remaining steps of the systematic review were completed on this software. A reference list of the included articles and reviews, as well as the cited articles of the relevant research, were investigated to find overlooked studies. Original longitudinal research articles (i) including patients diagnosed with MDD or BD according to the DSM criteria or ICD classification systems, (ii) including MDD or BD patients at an active depressive episode (unipolar or bipolar depression) and the follow-up of the same population, (iii) studies that reported mean and standard deviation of oxidative DNA damage changes before and after treatment of the depressive episode, (iv) with keywords or abstracts written in English, and (v) published from inception to 31.12.2024 were included in this review.

Screening and data extraction were performed by two independent reviewers through Covidence. A third reviewer resolved the conflicts during screening and cross-checked all the steps to prevent overlooked information. The authors of the included articles with missing data were contacted via email to request the data. The quality assessment tool on Covidence was customized according to the Newcastle-Ottawa Quality Assessment Scale to evaluate the risk of bias of the included studies in the meta-analysis.

### Statistical Analyses

Statistical analysis was performed on IBM SPSS Statistics 28.0 (Chicago IL, USA) for Windows. The effect sizes of the comparison between 8-OHdG levels before and after the treatment of the depressive episode were calculated by Hedges’s g estimation, using a random-effect model with 95% confidence intervals. I^2^ and Q tests were applied to evaluate the heterogeneity in the statistics. An I^2^ value equal to or more than 50% was set as a threshold of heterogeneity. Publication bias was assessed by employing Egger’s regression test and funnel plot. Subgroup analyses were conducted for unipolar and bipolar depression groups, pharmacological and medicine combined with ECT treatment, serum/plasma and urine samples, and measurement methods. Additionally, another meta-analysis was performed based on the data from only remitted patients. Meta-regression was also conducted based on sex (% female) by using effect sizes and standard errors (SE) of the meta-analyzed studies. The analysis was performed by including effect size as a dependent and sex percentage as an independent variable after cases were weighted according to inverse standard errors (1/SE).

## Results

In total 1512 studies were uploaded to Covidence, after duplicates were removed, 557 studies were screened. Out of 557, 25 articles were full-text screened and 15 of these weren’t eligible to be included in the systematic review. 9 out of 15 studies were excluded due to being reviews or meta-analyses, 2 were not a follow-up study [31, 32], 3 had the wrong patient population [33-35], and 1 didn’t measure 8-OHdG levels [36]. The remaining 10 studies were included in the systematic review and the meta-analyses. Figure 1 summarizes the literature search as a PRISMA flow diagram.

**Figure 1:**
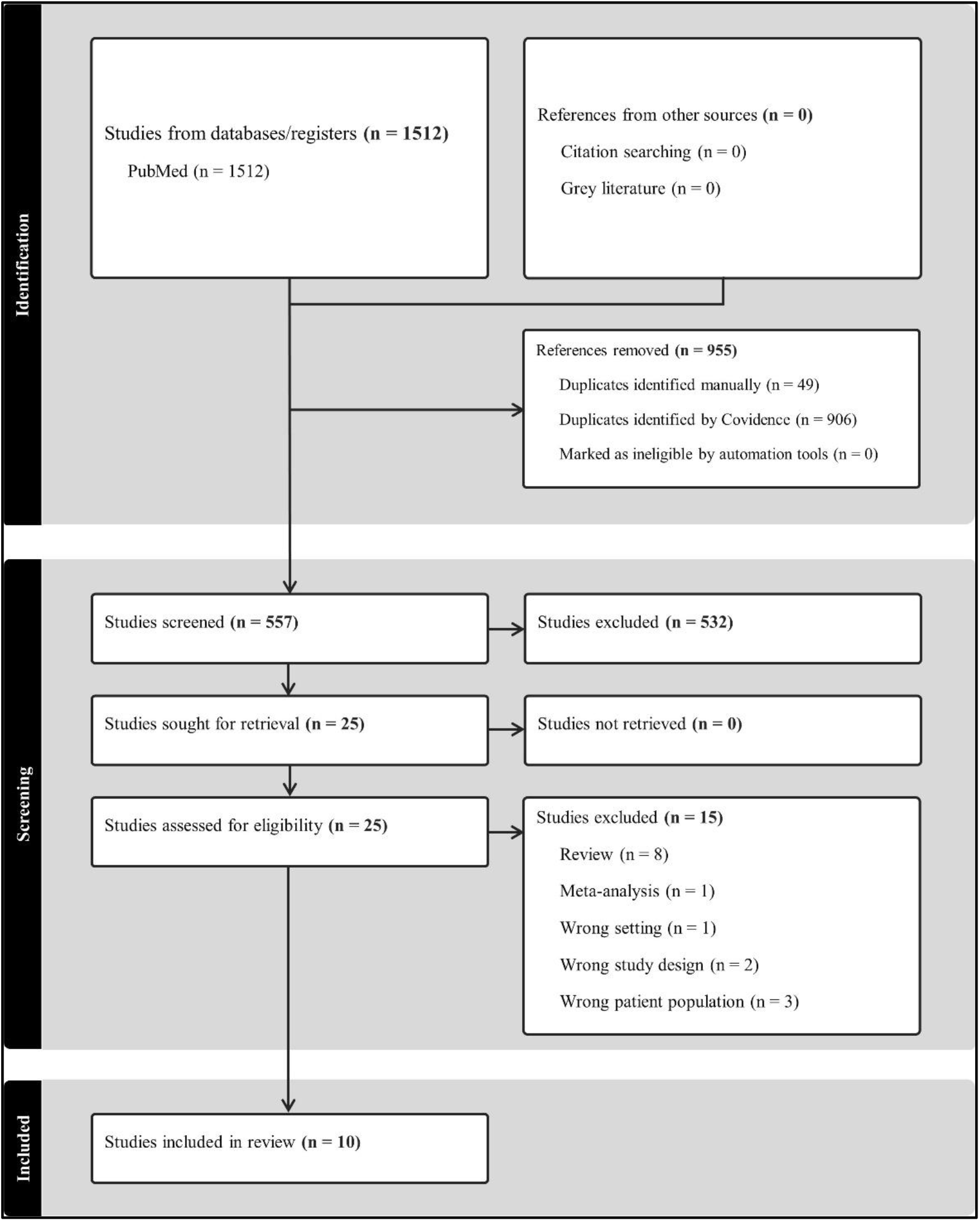
PRISMA flow diagram

Among the included studies, 5 investigated the 8-OHdG level change in UD [24, 37, 25, 26, 28], 1 studied only BD [27], and 4 examined both UD and BD populations [30, 28, 29, 38]. Additionally, of these studies, 8 applied pharmacological treatment [24, 37, 25, 26, 30, 27, 28, 38] while 2 combined medicine and electroconvulsive therapy (ECT) [39, 29]. There are also some variabilities in terms of biological sample sources and measurement methods. 6 studies collected blood samples [24, 37, 25, 26, 29, 38] while 3 obtained urine samples [39, 30, 28] and one studied with urine and salivary samples separately in the same population [27]. Moreover, in 6 articles, 8-OHdG levels were measured by enzyme-linked immunosorbent assay (ELISA) [24-27, 29, 38], 4 by liquid chromatography-tandem mass spectrometry (LC-MS/MS) [39, 37, 30, 28]. The change of 8-OHdG concentrations was either elevated [37, 27, 29], reduced [25, 26, 30, 27, 28] or didn’t change [39, 24, 37, 27, 38] after the follow-up. The results of the literature search are summarized in Table 1.

**Table 1:**
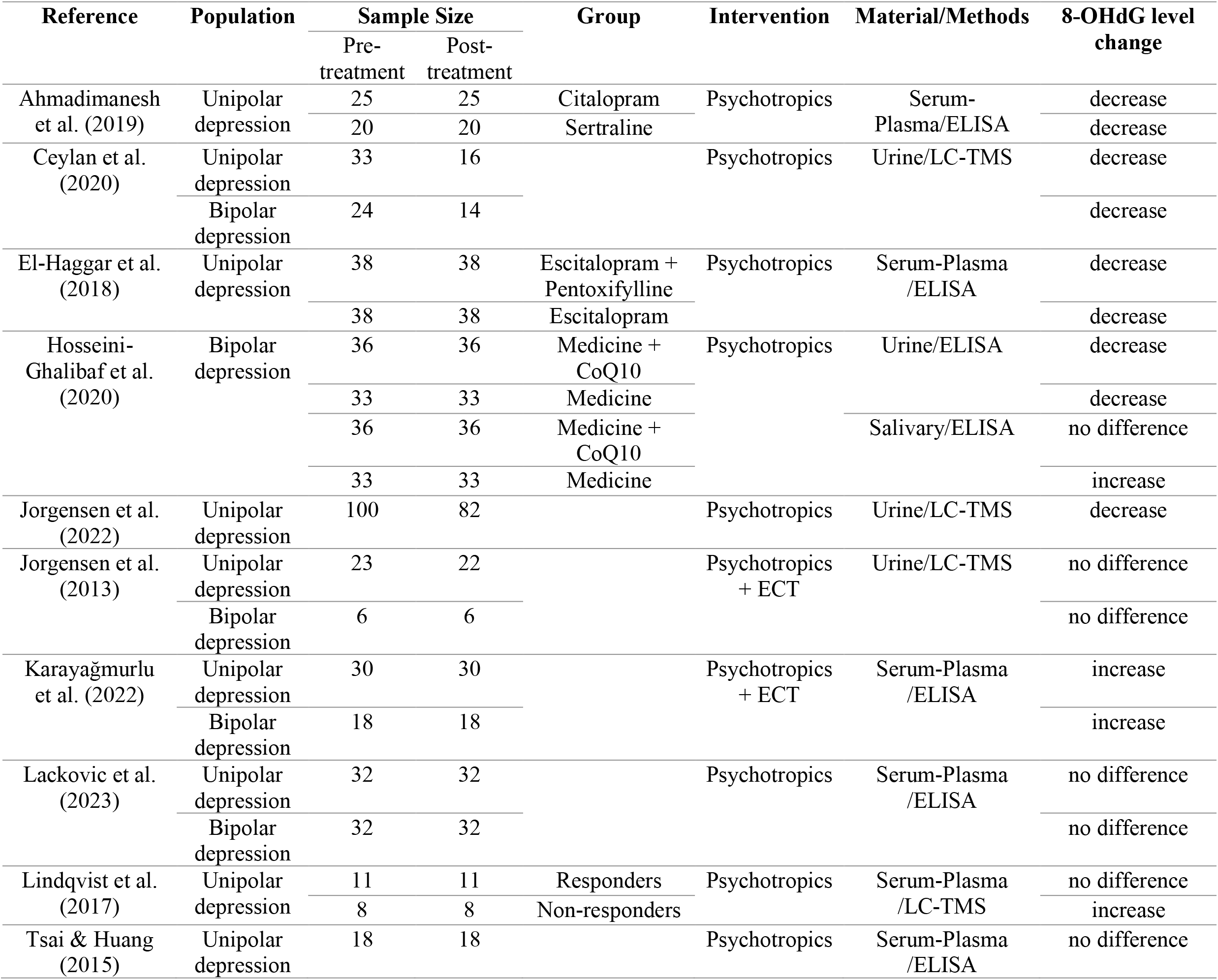
Studies that investigated the change of 8-OHdG levels after the intervention of an acute depressive episode.

### Follow-up of 8-OHdG levels after a depressive episode

In total, 525 individuals (UD=376, BD=149) were included in the meta-analysis for the pre-treatment, and 479 individuals (UD=340, BD=139) were followed up after treatment. As a result of the meta-analysis, 8-OHdG levels were significantly decreased after the treatment of the depressive episode (random effects: Hedge’s g = −0.61, 95% CI = −1.20 to −0.02, p = 0.04) (shown in Fig. 2). There was a significant heterogeneity across studies in the overall analysis (I^2^ = 0.95, Q = 207.45, df(Q) = 17, p < 0.001). There was no significant publication bias according to Egger’s regression-based Test (Coefficient = −0.246, 95% CI = −2.644 to 2.153, p = 0.831) (shown in Fig. 3).

**Figure 2:**
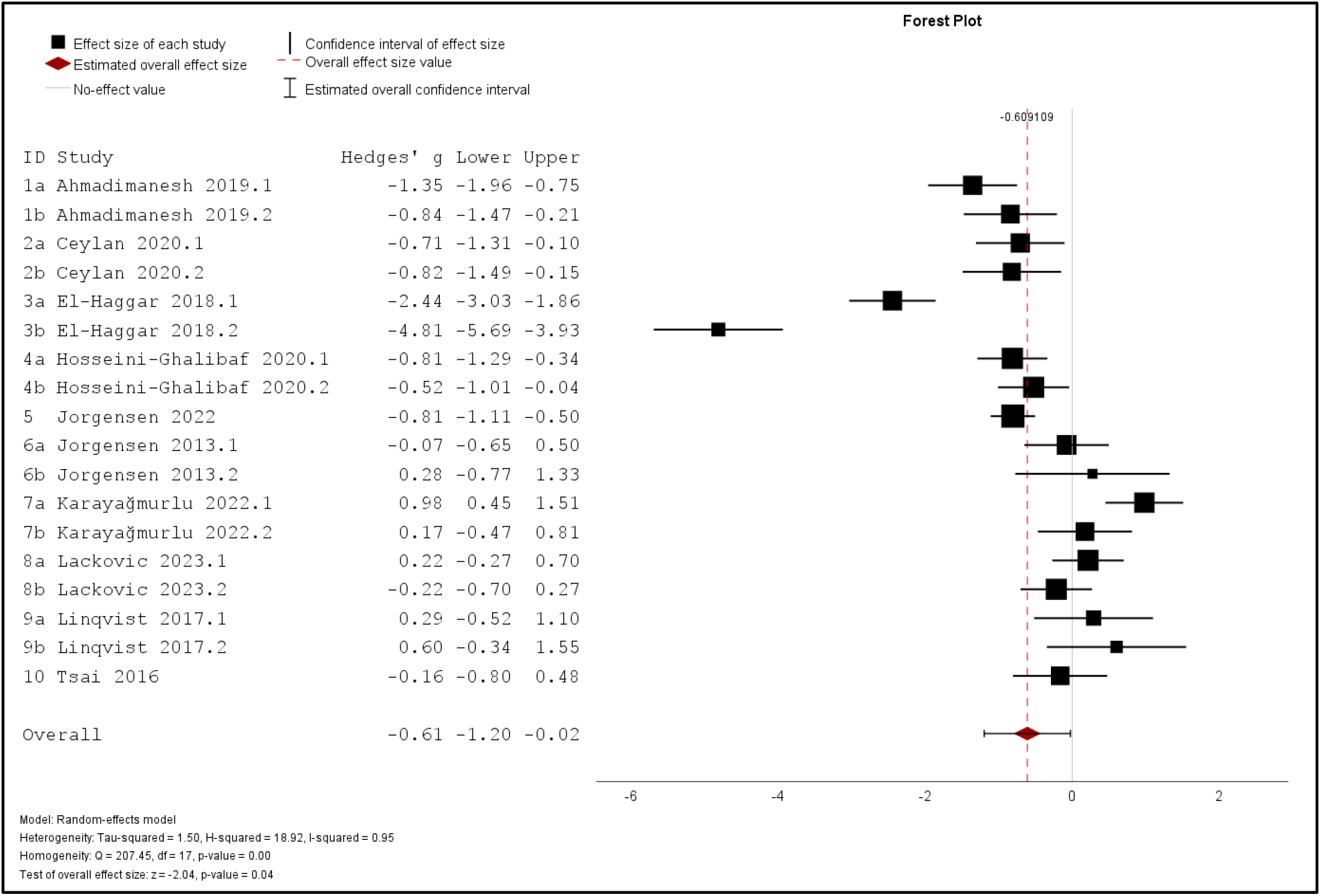
Meta-analysis of 8-OHdG levels change in unipolar and bipolar depression after intervention

**Figure 3:**
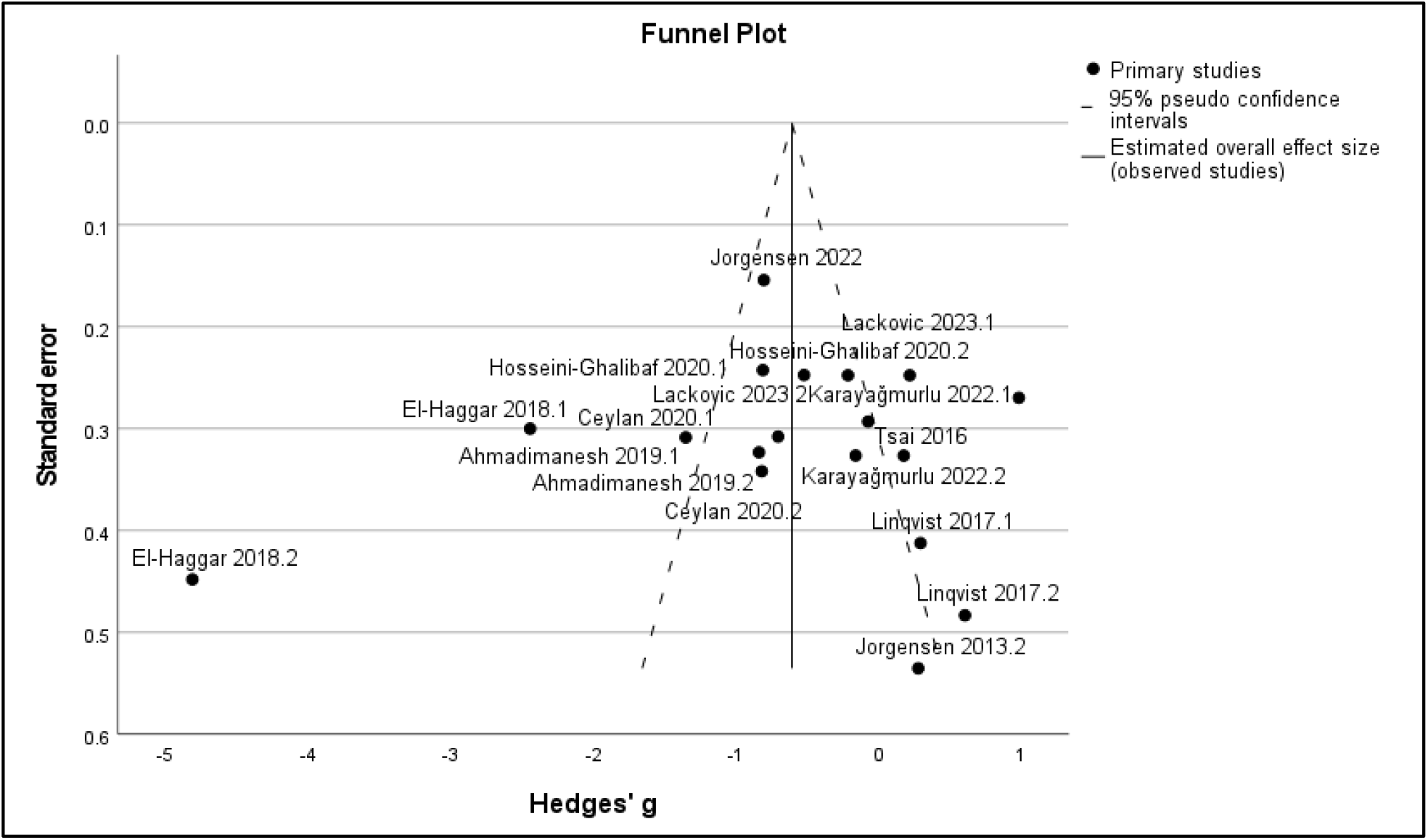
Funnel plot of the meta-analysis

### Follow-up of 8-OHdG levels in UD vs BD patients

Subgroup analyses were conducted for UD (9 studies, 12 groups) and BD (5 studies, 6 groups) patients. As a result, there was no significant reduction of 8-OHdG concentration in UD (random effects: Hedge’s g = − 0.75, 95% CI = −1.62 to 0.12, p= 0.09) while the BD group showed a significant decrease (random effects: Hedge’s g = −0.39, 95% CI = −0.73 to −0.05, p= 0.02). Significant heterogeneity was observed in the UD group (I^2^ = 0.96, Q = 195.432, df(Q) = 11, p < 0.001) whereas the BD group’s homogeneity was higher but close to the threshold (I^2^ = 0.49, Q = 9.853, df(Q) = 5, p = 0.080) (shown in Fig. 4). There was no significant publication bias according to the funnel plot (shown in Supplementary Fig. 1) Egger’s Regression-Based Test for both UD (Coefficient = 0.431, 95% CI = −3.531 to 4.393, p = 0.813) and BD groups (Coefficient = −1.147, 95% CI = −2.901 to 0.606, p = 0.143).

**Figure 4:**
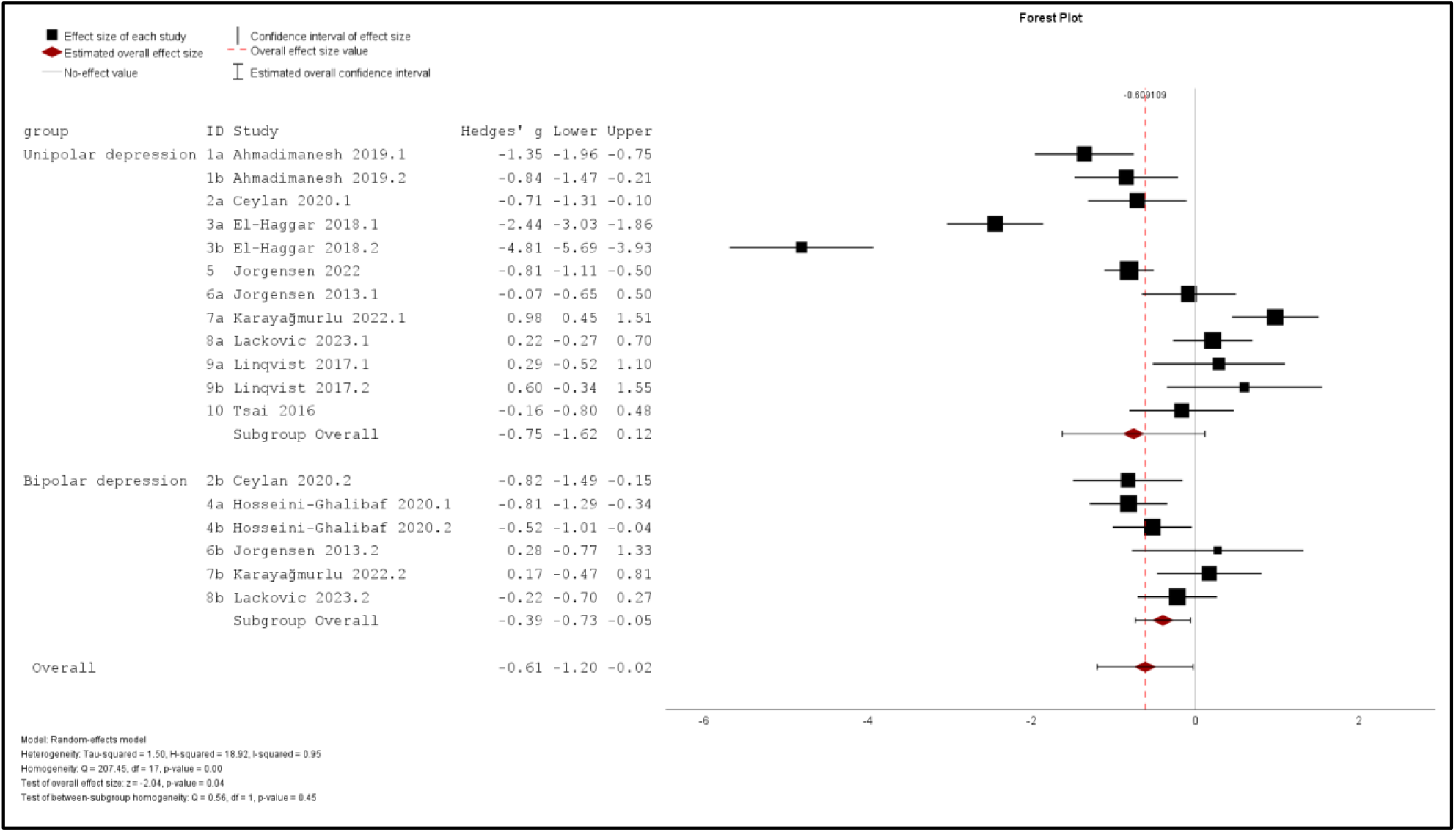
Subgroup analysis of 8-OHdG levels change after intervention in unipolar and bipolar depression

Additionally, to decrease the heterogeneity, subgroup analysis of UD (7 studies, 10 groups) and BD (3 studies, 4 groups) without ECT combined treatment were performed. There was a significant reduction of 8-OHdG concentration in UD (random effects: Hedge’s g = −1.00, 95% CI = −1.97 to −0.02, p= 0.04) as well as in the BD group (random effects: Hedge’s g = −0.57, 95% CI = −0.86 to −0.28, p < 0.001). However, heterogeneity wasn’t reduced in the UD group (I^2^ = 0.96, Q = 148.997, df(Q) = 9, p < 0.001) whereas the BD group’s homogeneity was elevated (I^2^ = 0.20, Q = 3.610, df(Q) = 3, p = 0.307) (shown in Supplementary Fig. 2). There was no significant publication bias according to Egger’s Regression-Based Test for both UD (Coefficient = −0.234, 95% CI = −4.738 to 4.270, p = 0.908) and BD groups (Coefficient = 0.162, 95% CI = −4.894 to 5.219, p = 0.903).

### Follow-up of 8-OHdG levels in pharmacological vs ECT combined treatment

Subgroup analyses were conducted for pharmacological intervention (8 studies, 14 groups) and pharmacological treatment combined with ECT (2 studies, 4 groups). There was a significant reduction of 8-OHdG concentration in pharmacological intervention (random effects: Hedge’s g = −0.88, 95% CI = −1.56 to −0.19, p = 0.001) while non-significant elevation was observed in ECT combined treatment (random effects: Hedge’s g = 0.36, 95% CI = −0.16 to 0.88, p= 0.17). Significant heterogeneity was observed in both pharmacological group (I^2^ = 0.95, Q = 156.271, df(Q) = 13, p < 0.001) and ECT combined group (I^2^ = 0.60, Q = 7.807, df(Q) = 3, p = 0.050). Forest plot and the funnel plot results of the analysis were shared as Supplementary Figure 3 and 4. There was no significant bias in publication according to Egger’s Regression-Based Test for pharmacological (Coefficient = 0.000, 95% CI = −2.969 to 2.970, p = 1.000) and ECT combined (Coefficient = 0.718, 95% CI = −4.035 to 5.471, p = 0.582) treatment groups.

### Follow-up of 8-OHdG levels in blood vs urine samples

Subgroup analyses were conducted regarding biological specimens: serum/plasma (6 studies, 11 groups) and urine (4 studies, 7 groups). As a result, there was no significant decrease of 8-OHdG concentration in serum/plasma samples (random effects: Hedge’s g = −0.68, 95% CI = −1.65 to 0.29, p= 0.17) while significance was observed in the studies with urine samples (random effects: Hedge’s g = −0.61, 95% CI = − 0.84 to −0.39, p< 0.001). Significant heterogeneity was observed in the studies that collected serum/plasma (I^2^ = 0.96, Q = 197.400, df(Q) = 10, p < 0.001) while there was a homogeneity in the urine samples (I^2^ = 0.24, Q = 8.854, df(Q) = 6, p = 0.182). Forest plot and the funnel plot results of the analysis were shared as Supplementary Figure 5 and 6. There was no significant publication bias according to Egger’s Regression-Based Test for serum/plasma (Coefficient = 1.269, 95% CI = −3.935 to 6.472, p = 0.595) whereas there was a bias for the urine group (Coefficient = −1.158, 95% CI = −1.892 to −0.424, p = 0.010).

### Follow-up of 8-OHdG levels in ELISA vs LC-MS/MS method

Subgroup analyses were conducted regarding the measurement method as ELISA (6 studies, 11 groups) and LC-MS/MS (4 studies, 7 groups). As a result, there was no significant decrease of 8-OHdG concentration in the measurement of ELISA (random effects: Hedge’s g = −0.87, 95% CI = −1.78 to 0.04, p= 0.06) and in the studies with LC-MS/MS measurement (random effects: Hedge’s g = −0.28, 95% CI = −0.71 to 0.15, p = 0.20). Significant heterogeneity was observed in the studies performed ELISA (I^2^ = 0.97, Q = 188.535, df(Q) = 10, p < 0.001) and the LC-MS/MS measure group (I^2^ = 0.68, Q = 18.367, df(Q) = 6, p = 0.005). Forest plot and the funnel plot results of the analysis were shared as Supplementary Figure 7 and 8. Publication bias according to Egger’s Regression-Based Test for the studies conducted ELISA (Coefficient = 4.826, 95% CI = 0.777 to 8.875, p = 0.025) was observed similarly with LC-MS/MS group (Coefficient = −1.389, 95% CI = −2.428 to −0.351, p = 0.018).

All subgroup analyses were summarized in Table 2.

**Table 2:** Summary of subgroup analyses.

| Subgroups | Number of analyzed groups | Hedges' $g$ (95% CI) | $I^2$ | Q-test | Q-test $p$ | Bias (95% CI) |
| --- | --- | --- | --- | --- | --- | --- |
| **Depression type** |  |  |  |  |  |  |
| Unipolar depression | 12 | 0.75 (-1.62 to 0.12) | 0.96 | 195.432 | <0.001 | 0.431 (-3.531 to 4.393) |
| Bipolar depression | 6 | -0.39 (-0.73 to -0.05) | 0.49 | 9.853 | 0.080 | -1.147 (-2.901 to 0.606) |
| **Treatment method** |  |  |  |  |  |  |
| Pharmacological | 14 | -0.88 (-1.56 to -0.19) | 0.95 | 156.271 | <0.001 | 0.000 (-2.969 to 2.970) |
| ECT combined | 4 | 0.36 (-0.16 to 0.88) | 0.60 | 7.807 | 0.05 | 0.718 (-4.035 to 5.471) |
| **Biological specimen** |  |  |  |  |  |  |
| Serum/Plasma | 11 | -0.68 (-1.65 to 0.29) | 0.96 | 197.400 | <0.001 | 1.269 (-3.935 to 6.472) |
| Urine | 7 | -0.61 (-0.84 to -0.39) | 0.24 | 8.854 | 0.182 | -1.158 (-1.892 to -0.424) |
| **Measurement method** |  |  |  |  |  |  |
| ELISA | 11 | -0.87 (-1.78 to 0.04) | 0.97 | 188.535 | <0.001 | 4.826 (0.777 to 8.875) |
| LC-MS/MS | 7 | -0.28 (-0.71 to 0.15) | 0.68 | 18.367 | 0.005 | -1.389 (-2.428 to -0.351) |

### Changes of 8-OHdG levels after reaching remission

Another dataset was prepared with the data from only remitted patients, which was shared in 3 studies [30, 28, 38] and 5 different groups. The characteristics of the studies, including remission status, remission criteria, and follow-up duration were presented in Supplementary Table 1. In total, 221 individuals (UD=165, BD=56) were included in the meta-analysis for the pre-treatment, and 117 individuals (UD=71, BD=46) were followed up after treatment. As a result of the meta-analysis, 8-OHdG levels were significantly decreased after the treatment of the depressive episode (random effects: Hedge’s g = −0.42, 95% CI = −0.81 to −0.03, p= 0.03). There was a significant heterogeneity across studies in the overall analysis (I^2^ = 0.63, Q = 10.91, df(Q) = 4, p = 0.03). The forest plot and the funnel plot results of the analysis were shared as supplementary material. There was no significant publication bias according to Egger’s Regression-Based Test (Coefficient = 1.006, 95% CI = −3.307 to 5.319, p = 0.512).

### Meta-regression based on sex

To examine the potential moderating effect of sex on oxidative DNA damage after follow-up, a meta-regression analysis was conducted. This analysis displayed that sex was not significant as a predictor of 8-OHdG changes in the overall population (β = 0.018, 95% CI [−0.006, 0.42], p = 0.147). However, when the same analysis was performed in terms of UD and BD, UD indicated significant results (β = 0.099, 95% CI [0.065, 0.133], p < 0.001), while sex is not an 8-OHdG change mediator on BD (β = 0.017, 95% CI [−0.001, 0.036], p = 0.066).

## Discussion

In this study, existing literature about the follow-up of oxidative DNA damage marker 8-OHdG after treatment of depressive episodes was systematically reviewed. Ten longitudinal studies were reviewed, and 10 were included in the meta-analysis with 376 UD and 149 BD patients. Regardless of limited literature, variability in methodology, sample sizes, and clinical features, this study demonstrated that 8-OHdG levels declined in UD and BD patients after the follow-up. Additionally, this meta-analysis showed that even though the patients couldn’t reach a stable phase until the follow-up in most of the studies, 8-OHdG concentration tends to diminish. On the other hand, the second meta-analysis with 165 remitted UD and 56 euthymic BD patients indicated similar results.

Most studies investigating oxidative DNA damage in a depressive state measured 8-OHdG levels, one of the main DNA oxidation products, to evaluate DNA damage [40, 33, 23]. Quantitative measurements of oxidative DNA damage in depression have been found to be increased consistently in previous studies with various biological specimens, such as blood and urine, and different measurement methods such as immunoassays and chromatographic techniques [40]. In this study, we investigated how 8-OHdG levels change after an intervention of a depressive episode to understand its episodic value as a biomarker by taking all the main variables into account that might be interfering with the results.

In the literature, 8 studies investigated the 8-OHdG level changes after intervention in the UD patient cohort. 2 of these studies demonstrated similar results, decreased levels of 8-OHdG from the blood samples after pharmacological therapy [25, 26], while Tsai & Huang (2015) found non-significant reduction [24] and Lackovic et al. (2023) obtained non-significant elevation of oxidative DNA damage level with similar sample collection and intervention method [38]. Moreover, the study conducted by Lindqvist et al. (2017) found that the population with UD patients who didn’t respond to pharmacological treatment had increased 8-OHdG levels. In contrast, there were no significant differences in the responded patient group [37]. On the other hand, from urine samples, Ceylan et al. (2020) and Jorgensen et al. (2022) showed a reduced concentration of 8-OHdG after pharmacological intervention [30, 28]. Besides, the study by Karayağmurlu et al. (2022) combined pharmaceutical therapy with ECT as a treatment and obtained an increased level of oxidative DNA damage in blood samples from UD patients [29]. Additionally, a study with a cohort of UD and BD found non-significant results, such as non-significant reduction in UD and non-significant elevation of 8-OHdG levels in BD after ECT treatment with medicine therapy in urinary samples [39].

Moreover, 4 studies examined similar factors in the BD cohort [30, 27, 29, 38]. Among these, Ceylan et al. (2020) from urine samples obtained reduced levels of 8-OHdG after medicine treatment [30] while Lackovic et al. (2023) from blood samples found similar but non-significant results [38]. Karayağmurlu et al. (2022) employed a combination of medicine and ECT as a treatment of BD and found raised 8-OHdG concentrations from blood [29]. Moreover, a study held by Hosseini-Ghalibaf et al. in 2020 collected urinary and salivary samples to investigate the effect of coenzyme Q10 supplementation along with pharmaceutical therapy in BD patients. They obtained various results depending on the sample; 8-OHdG levels were decreased in the urine samples while there was either no significant difference or increase in the samples from salivary [27].

Ten out of 10 eligible studies were included in the meta-analysis. Firstly, overall meta-analysis was conducted with 18 different populations from 10 studies. This analysis showed a significant decrease in oxidative DNA damage after the intervention of a depressive episode with great heterogeneity. Considering this analysis consisted of both UD and BD patients with different treatment methods, biological specimens, and laboratory techniques, heterogeneity was anticipated. In order to decrease the heterogeneity, subgroup analyses were performed for each parameter.

According to subgroup analyses, UD patient populations showed no significant change in oxidative DNA damage with high heterogeneity, while in BD population, not only the level of 8-OHdG was significantly decreased but also homogeneity across studies were observed. Even though homogeneity is very close to the threshold, heterogeneity was lower in BD studies compared to the UD group, which can be explained by the differential disease pathogenesis. To increase homogeneity in both groups, another meta-analysis was conducted by excluding the studies with ECT combined treatment. This resulted in significant reduction in oxidative DNA damage in UD population with a high heterogeneity while in BD, not only the significant decrease was maintained but also homogeneity was elevated. Both of these diseases have heterogeneous features in terms of neuropathological and etiological mechanisms [41, 42]. However, BD shows higher homogeneity in contrast to UD through its consistent onset patterns and distinct clinical presentation with a greater genetic component [43].

In addition, intervention methods were compared by another subgroup analysis. The number of studies employed ECT with medicine treatment was very few but the number of the groups inside these studies was enough for meta-analysis. This analysis indicated that pharmacological therapy has significantly decreased the 8-OHdG level while ECT combined therapy has contrasting but non-significant effects by increasing oxidative DNA damage. Among most of these studies, the number of patients that have reached remission state after the pharmacological treatment is very limited as summarized in Supplementary Table 1, which might suggest that the reduced level of 8-OHdG might be due to medication’s antioxidant effects rather than recovering from the depressive episode. The antioxidant effect of the antidepressants as a treatment of UD was found to be associated with decreased oxidative DNA damage previously [23]. On the other hand, when medication was combined with ECT, the effect varies by contrasting results. ECT might cause greater oxidative stress through electrical fields or by induced seizures that can’t compensate for the antioxidant effects of the medication. However, according to a systematic review investigating the oxidative stress and ECT relation, there is not enough data to support this relationship with inconsistent findings [44]. Moreover, it should be noted that the results of this subgroup analysis are dependent on only 2 studies with small sample sizes. Thus, more studies are needed with ECT treatment to show this association.

Next analysis was held regarding the biological specimens as being serum/plasma from blood or urine. Although there was one study with saliva and urine samples from the same population, since there were no other studies with saliva samples, only the data coming from the urine samples were included in the analysis [27]. No significant change was observed with the blood samples, while there was a significant decrease of 8-OHdG in urine samples. Moreover, the homogeneity was high among the studies with urine samples, as is the opposite with blood specimens. A previous meta-analysis held by Goh et al. (2021) on oxidative DNA damage in UD, BD, and schizophrenia patients also indicated significant results in BD population when the studies with urine samples were analyzed. Similarly, no significant findings were reported with blood samples in any of the groups [14]. Not only the findings of this meta-analysis but also the study conducted by Hosseini-Ghalibaf et al. (2020) showed the importance of the biological specimen while detecting 8-OHdG concentrations [27, 14]. According to this study, results of the oxidative DNA damage levels from the same BD patient population differed based on the specimen type. The 8-OHdG concentration was significantly decreased in urine samples whereas there was an increase in salivary samples when patients were treated with only medication [27]. These differences might be observed due to 8-OHdG metabolism.

With oxidative DNA damage, 8-OHdG production occurs and is released into the bloodstream. Later, it is filtered through kidneys and excreted in the urine. That’s why the concentration of 8-OHdG is affected by blood clearance. However, urinary samples are not affected by these types of mechanisms; in contrast, they could be a better specimen candidate to measure overall oxidative DNA damage coming from multiple tissues [45, 46]. In addition, a previous study revealed that urinary 8-OHdG levels are more stable compared to those with blood samples, which makes urine a more reliable candidate of oxidative DNA damage source [47].

Comparison across measurement methods was also performed through another subgroup analysis. In all included studies, either immunoassays such as ELISA or chromatographic methods such as LC-MS/MS were used to detect the concentrations of 8-OHdG. The analysis revealed no significant changes when the measurement was performed by ELISA or the LC-MS/MS method. Similarly, the heterogeneity was very high in both groups. ELISA is an effective, reliable, and highly sensitive measurement method that can be applied to both blood and urine samples [48, 49]. LC-MS/MS is also a sensitive and selective quantifying technique, especially from plasma and urine, which is employed commonly across studies [50, 45, 23]. According to a previous meta-analysis that was conducted by Black et al. (2015), the oxidative stress level was higher in depression when measured with immuno-assays compared to chromatographic methods [20]. However, the correlation between these two measures is generally high [51], suggesting the interfering effect on the association with depression is low [20]. On the other hand, as discussed in the same study, it should also be noted that the studies collected urine samples mainly chose chromatographic measures while the rest with blood samples performed quantification by immuno-assays. Thus, instead of the measurement technique, biological specimens could be the main underlying reason for this difference [20].

Lastly, another data set was prepared with the data only coming from remitted patients. 2 studies were already conducted with only follow-up of remitted patients [30, 38] while the study by Jorgensen et al. (2022) shared the mixture of remitted and non-remitted patients at the same time [28]. However, the needed data were extracted through shared graphs and included in the meta-analysis. Since there wasn’t any available data regarding remitted patients in other studies, only 165 UD and 56 BD from 3 studies with 5 different groups were analyzed. As a result, there was a significant reduction in oxidative DNA damage with heterogeneity. This analysis was conducted to investigate the effects of actual recovery after the follow-up period. However, since all these studies intervene in the depressive episode by pharmacological treatment, it’s not possible to point out that the decrease was caused due to remission. In addition, these studies have conflicting results with high heterogeneity and small sample sizes; more studies are needed to understand the potential role of 8-OHdG as an episodic biomarker.

Besides meta-analysis, meta-regression analysis was also performed for the overall and subpopulations of UD and BD. As a result of this analysis, it was revealed that sex was not a significant predictor of oxidative DNA damage change not only in the overall population but also in the BD subpopulation. In contrast, the significance in the UD population indicated that the female sex contributes to variations in 8-OHdG levels during follow-up. However, it’s noteworthy that BD population almost reached statistically significant results, which might suggest statistical power limitations due to the relatively small number of BD population. In the literature, investigating sex differences in terms of oxidative DNA damage in affective disorders has indicated that females may be more vulnerable to elevated oxidative stress as a response to depression [52-54, 35], while there is no observed relationship in males [53]. Moreover, in BD, previous studies reported sex-specific clinical features and biological markers [55, 10] along with some findings showing higher oxidative stress levels in female BD patients [49]. Therefore, the association of sex and illness is likely to exist for the BD population as well and can be uncovered with future studies that have larger sample sizes [56].

Our findings contribute to understanding the treatment of affective disorders from a biological perspective through oxidative DNA damage. Previous meta-analysis has already presented that 8-OHdG could be an episode-related marker rather than a trait marker [22]. However, our study indicates that 8-OHdG level change might also be a valuable marker to evaluate the treatment response and recovery progression. This study has many indications in terms of understanding the change in oxidative DNA damage in depressive populations in a longitudinal period. However, there are also many limitations. Firstly, the sample size of the meta-analysis is limited, and high heterogeneity was observed in most of the analyses, which indicates there is great variability across studies. Even with subgroup analysis aiming to categorize similar features together, it didn’t diminish the heterogeneity level due to having many parameters at the same time. Secondly, some of the studies didn’t share the descriptive data in the article; instead, they preferred to show the findings by using graphs. For these types of studies, we extracted data approximately from the graphs, which might end up with minor errors. Thirdly, the confounding factors that might interfere with the oxidative DNA damage, such as smoking, obesity, lifestyle, and type of medication, differ across studies, and we didn’t involve these in the meta-analysis. Fourthly, since there weren’t enough studies on different kinds of treatment methods, this meta-analysis mainly focused on pharmaceutical interventions. Finally, in the studies included, most of the patients weren’t remitted until the follow-up period, and the data of the remitted and non-remitted patients haven’t been mentioned separately, which caused a variety across studies as well.

The results of this systematic review and meta-analysis indicate that the level of oxidative DNA damage marker, 8-OHdG, decreases after an intervention of a depressive episode in MDD and BD, not only in the overall population of both responders and remitters but also in remitted patients when analysed alone. The 8-OHdG concentration can be a potential biomarker for treatment response particularly in BD. Additionally, 8-OHdG appears to have a more consistent performance as a biomarker for treatment with pharmacological agents than ECT. Findings point at urine as a more convenient biosample for detection of changes in 8-OHdG levels with treatment. For unipolar depression, sex should be taken into consideration while studying changes in oxidatively induced DNA damage with treatment as the female sex was found to be a significant moderator of this change in response to treatment in the UD population. Further longitudinal studies using ECT combined treatment, urine sampling, and more remitted patients with longer duration of follow-up are needed to evaluate treatment-related changes in oxidatively induced damage in depression.

## Statements

## Acknowledgement

The authors gratefully acknowledge the use of the services and facilities of the Koç University Research Center for Translational Medicine (KUTTAM), funded by the Presidency of Turkey, Head of Strategy and Budget. Additionally, the authors acknowledge the assistance of a librarian from Koç University, Suna Kiraç Library.

## Statement of Ethics

Ethics approval was not required for this study.

## Conflict of Interest Statement

The authors have no conflicts of interest to declare.

## Funding Sources

This research was funded by Koç University Research Center for Translational Medicine (KUTTAM). The funder had no role in the design, data collection, data analysis, and reporting of this study.

## Author Contributions

Buket Yeşiloğlu: data curation, methodology, formal analysis, investigation, resources, project administration, writing – original draft, and writing – review and editing. Ceren Gör: data curation, writing – original draft, and writing – review and editing. Reyhan Nur Babalıoğlu: data curation, writing – review and editing. şevin Hun şenol: data curation, writing – review and editing. Mark Frye: writing – review and editing, and supervision. Anders Jørgensen: writing – review and editing, and supervision. Ayşegül Özerdem: writing – review and editing, and supervision. Deniz Ceylan: conceptualization, data curation, methodology, investigation, resources, project administration, writing – original draft, writing – review and editing, and supervision.

## Data Availability Statement

All data generated or analyzed during this study are included in this article and its supplementary material files. Further enquiries can be directed at the corresponding author.

## Notes

### Competing Interest Statement

The authors have declared no competing interest.

### Clinical Protocols

https://www.crd.york.ac.uk/PROSPERO/view/CRD42025630675

### Funding Statement

This research was funded by Koc University Research Center for Translational Medicine (KUTTAM).
The funder had no role in the design, data collection, data analysis, and reporting of this study.

