## Supplemental Materials for "Oxidative DNA Damage Diminishes After Treatment of Unipolar and Bipolar Depressive Episodes: A Systematic Review and Meta-Analysis of Longitudinal Studies"

**Supplementary Materials**

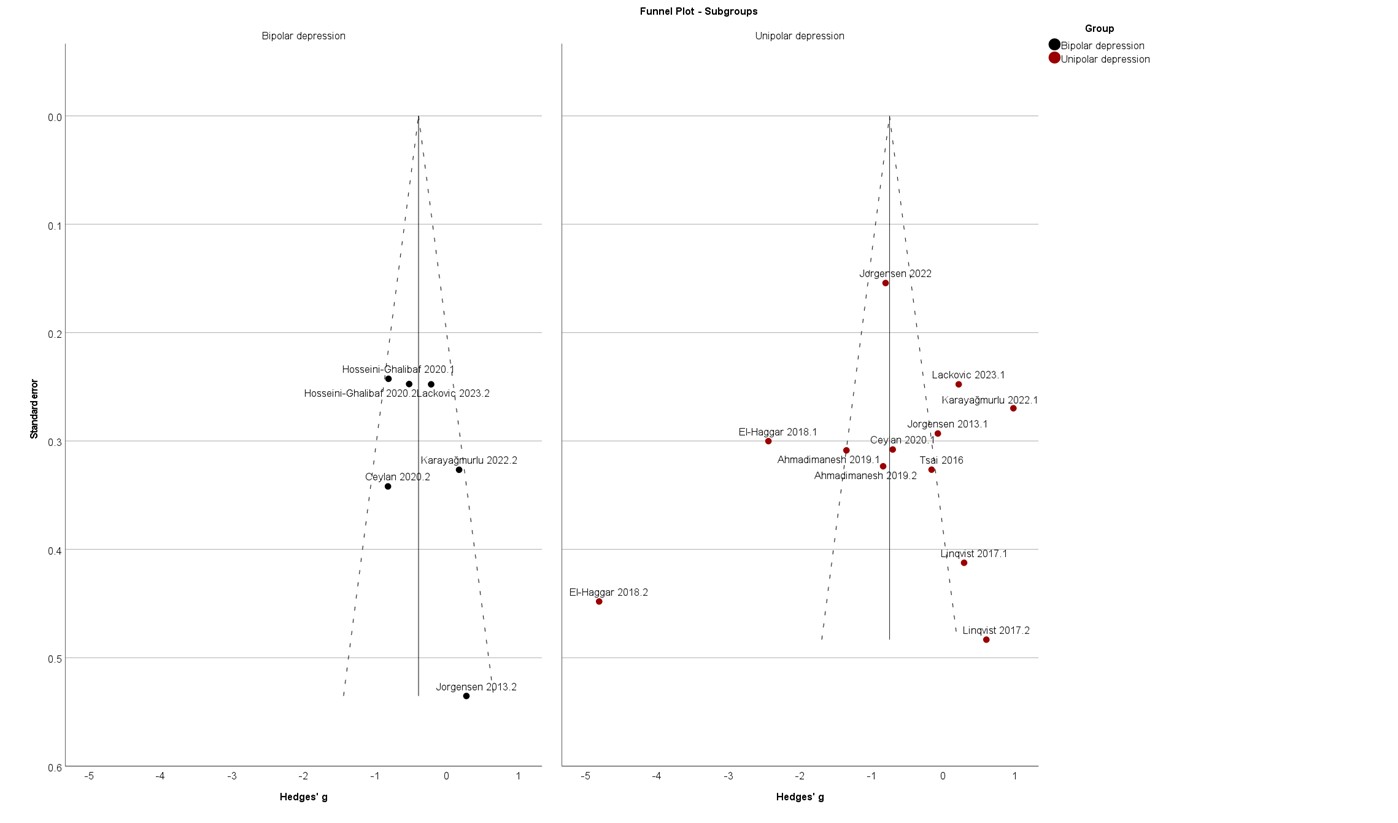

Figure 1: Funnel plot of the subgroup analysis of 8-OHdG levels change after intervention in unipolar and bipolar depression

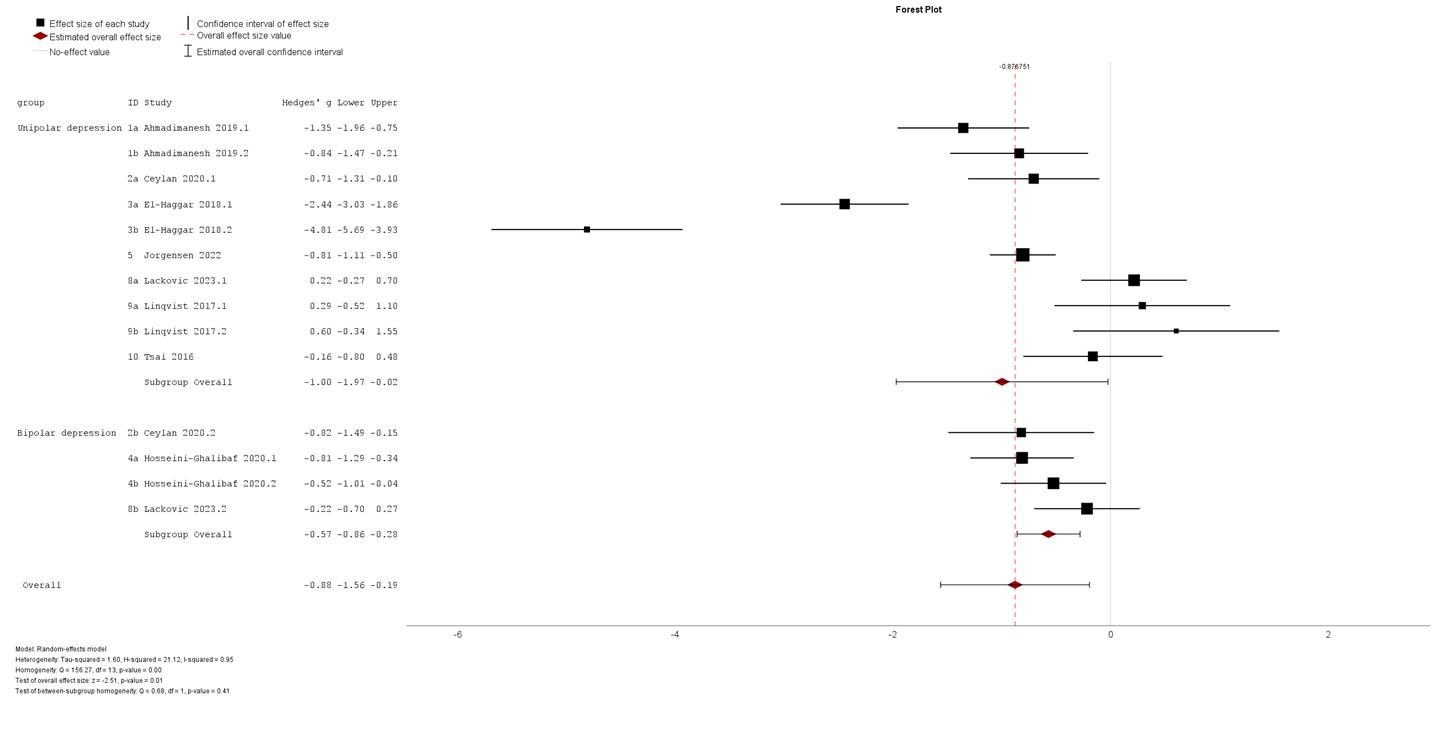

Figure 2: Subgroup analysis of 8-OHdG levels change after pharmacological intervention in unipolar and bipolar depression

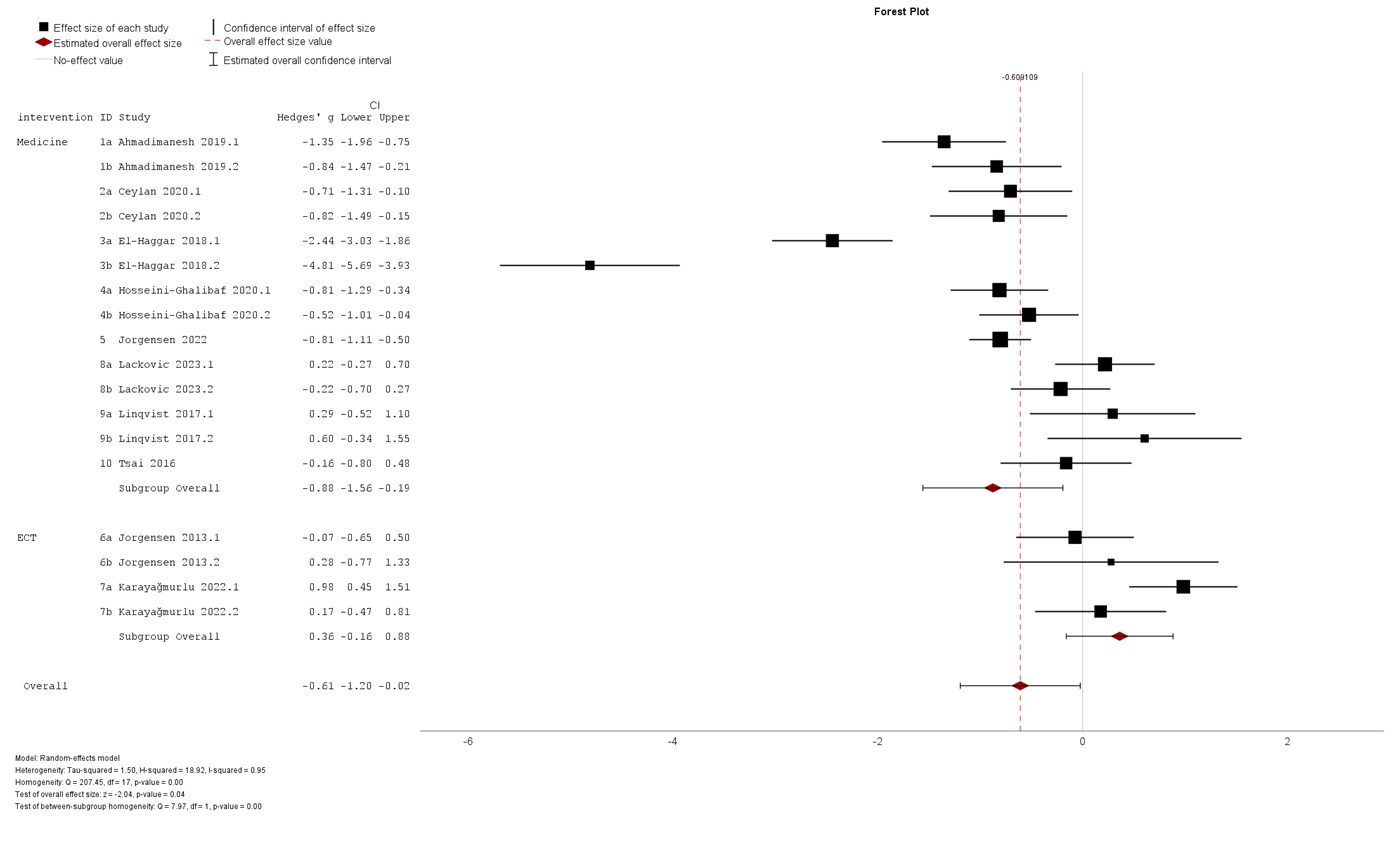

Figure 3: Subgroup analysis of 8-OHdG levels change after pharmacological and ECT combined intervention

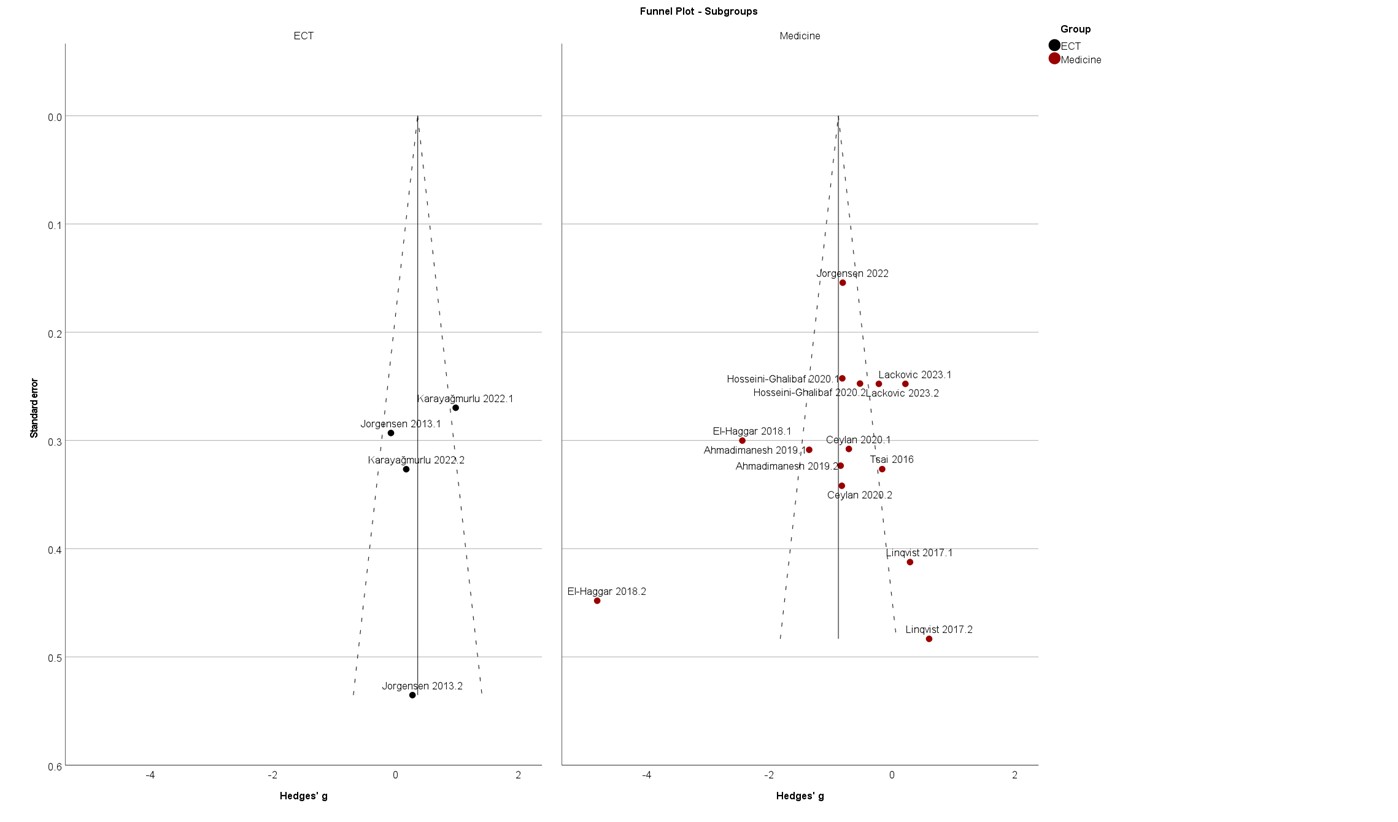

Figure 4: Funnel plot of the subgroup analysis of 8-OHdG levels change after pharmacological and ECT combined intervention

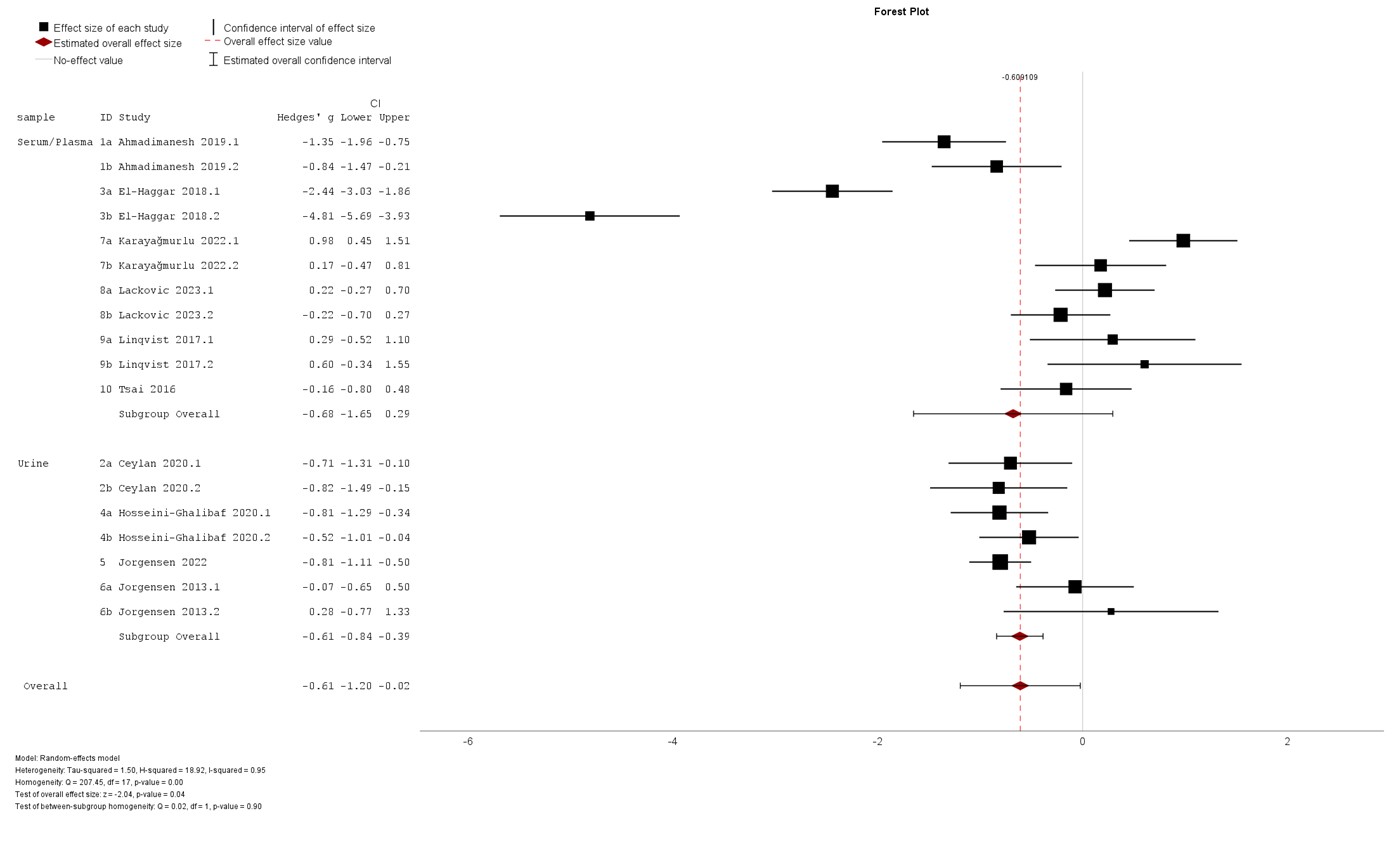

Figure 5: Subgroup analysis of 8-OHdG levels change of serum/plasma and urine sampling

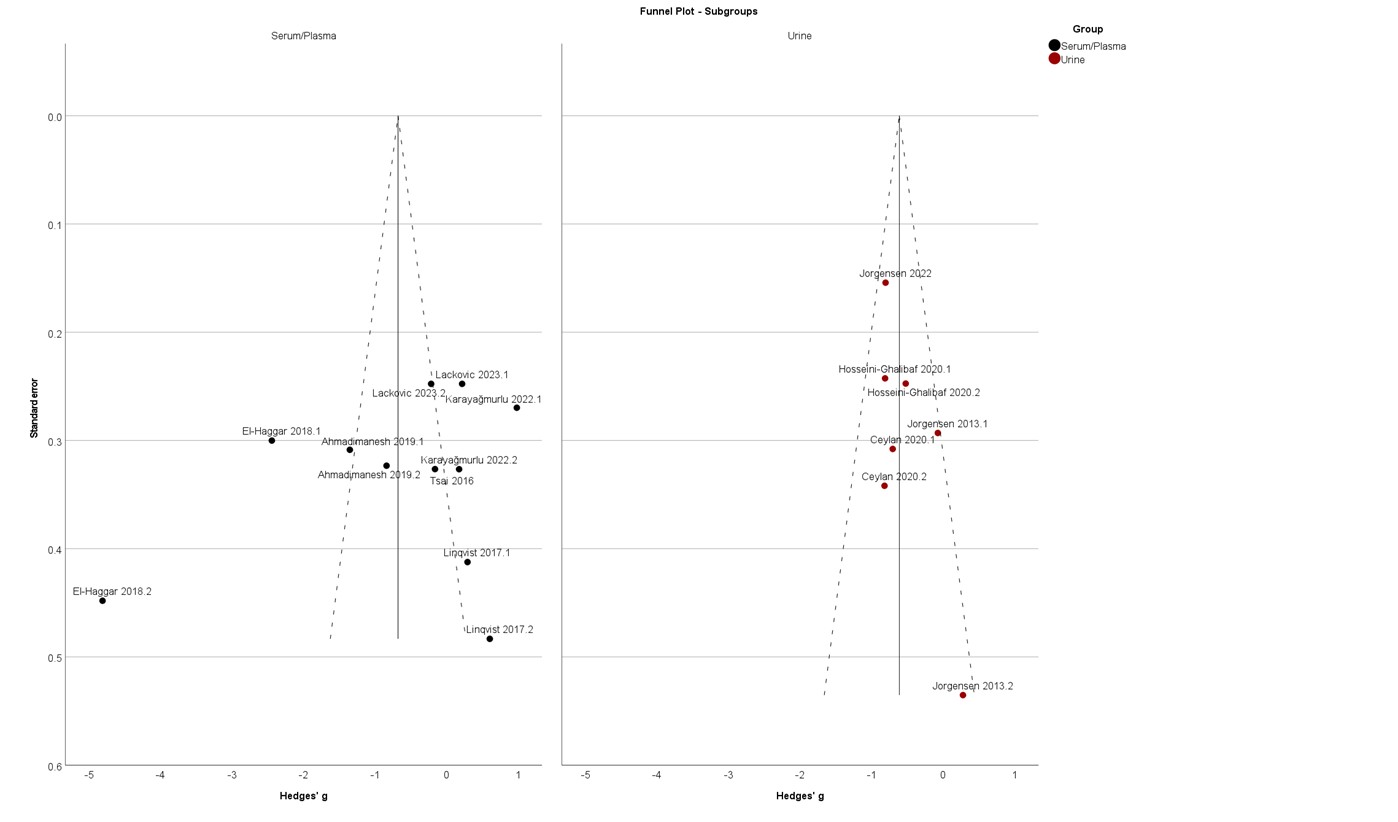

Figure 6: Funnel plot of the subgroup analysis of 8-OHdG levels change of serum/plasma and urine sampling

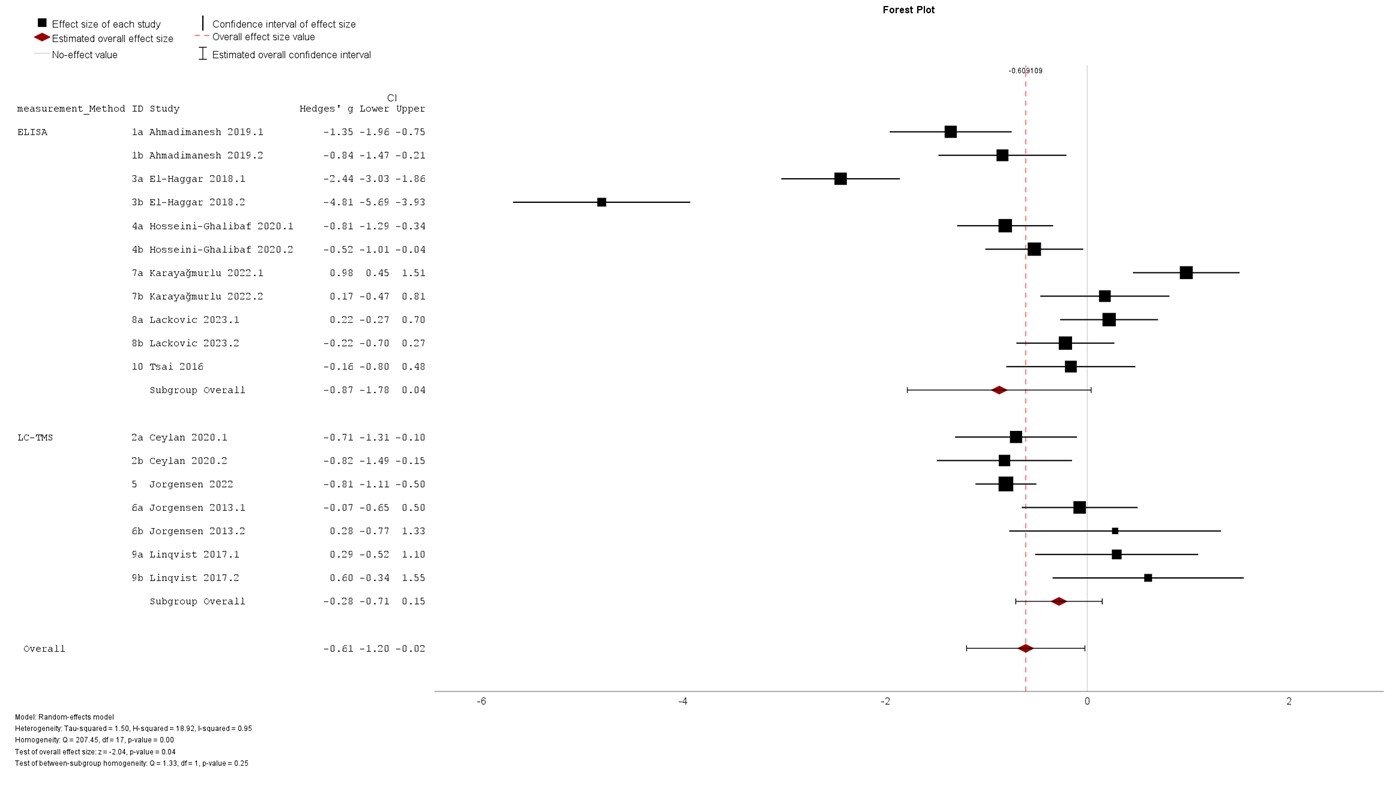

Figure 7: Subgroup analysis of 8-OHdG levels change of ELISA and LC-MS/MS methods

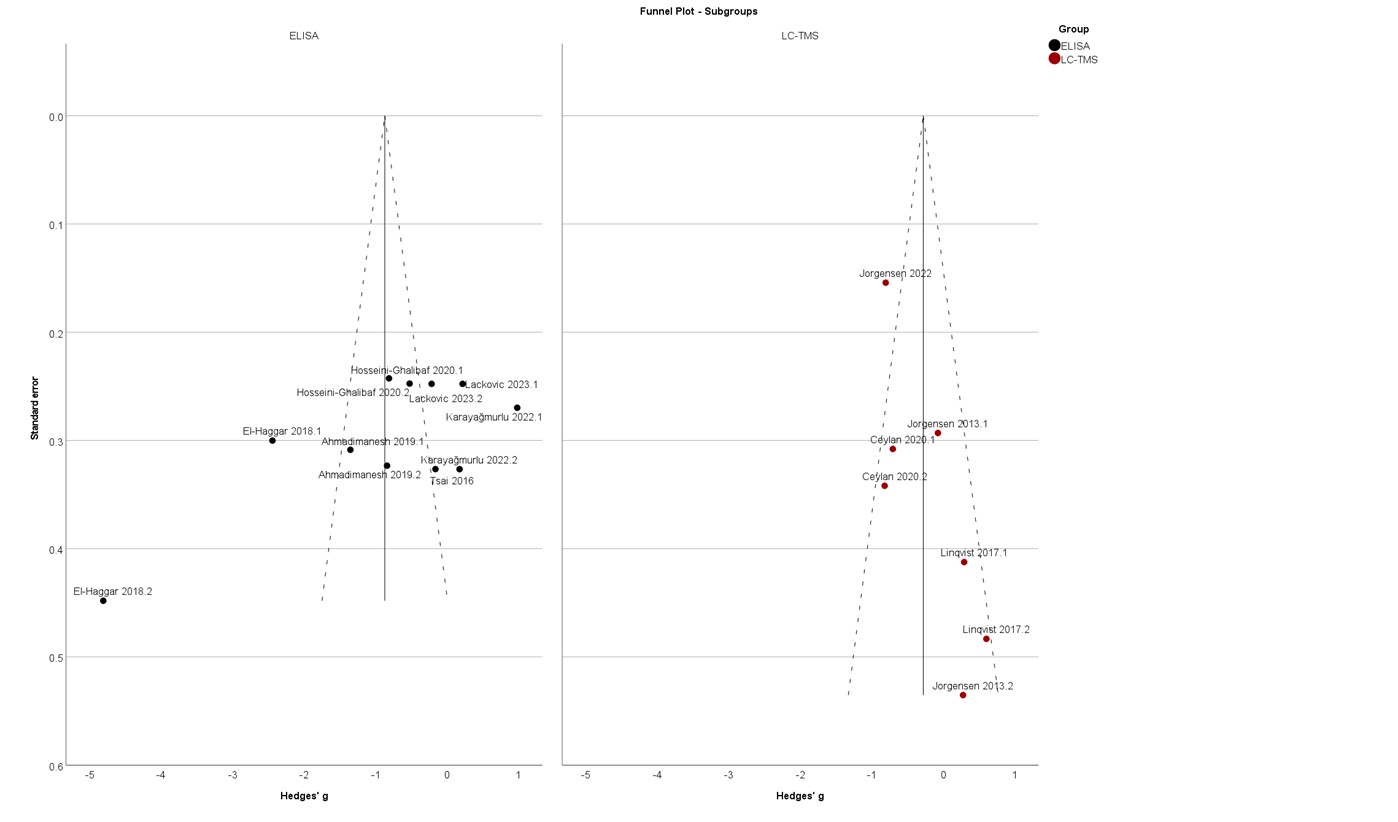

Figure 8: Funnel plot of the subgroup analysis of 8-OHdG levels change of ELISA and LC-MS/MS methods

Table 1: Detailed summary of the included studies with remission data

| Reference | Population | Group | Sample Size | | Number of remitted patients | Remission criteria | Follow-up duration | Intervention | 8-OHdG level change |
| --- | --- | --- | --- | --- | --- | --- | --- | --- | --- |
|  |  |  | Pre-treatment | Post-treatment |  |  |  |  |  |
| Ahmadimanesh et al. (2019) | Unipolar depression | Citalopram | 25 | 25 | 5 | HDRS-17 scores≤7 | 15 weeks | Psychotropics | decrease |
|  |  | Sertraline | 20 | 20 | 6 |  |  |  | decrease |
| **Ceylan et al. (2020)** | Unipolar depression |  | 33 | 16 | 16 | not meeting the diagnostic criteria in the SCID-I HDRS-17 scores<7 | 8 weeks | Psychotropics | decrease |
|  | Bipolar depression |  | 24 | 14 | 14 |  |  |  | decrease |
| El-Haggar et al. (2018) | Unipolar depression | Escitalopram + Pentoxifylline | 38 | 38 | 32 | HDRS-17 scores≤7 | 12 weeks | Psychotropics | decrease |
|  |  | Escitalopram | 38 | 38 | 16 |  |  |  | decrease |
| Hosseini-Ghalibaf et al. (2020) | Bipolar depression | Medicine + CoQ10 | 36 | 36 | 3 | MADRS ≤7 | 8 weeks | Psychotropics | decrease |
|  |  | Medicine | 33 | 33 | 0 |  |  |  | decrease |
| **Jorgensen et al. (2022)** | Unipolar depression |  | 100 | 82 | 23 | ≥50% reduction in HDRS-6 at week 4 and HDRS-6 score <5 at week 8 | 8 weeks | Psychotropics | decrease |
| Jorgensen et al. (2013) | Unipolar depression |  | 23 | 22 | 9 | HDRS-17 scores≤7 | 1 week post-ECT evaluation | Psychotropics + ECT | no difference |
|  | Bipolar depression |  | 6 | 6 |  |  |  |  | no difference |
| Karayağmurlu et al. (2022) | Unipolar depression |  | 30 | 30 | NA | NA | NA | Psychotropics + ECT | increase |
|  | Bipolar depression |  | 18 | 18 | NA |  |  |  | increase |
| **Lackovic et al. (2023)** | Unipolar depression |  | 32 | 32 | 32 | NA | 10 weeks | Psychotropics | no difference |
|  | Bipolar depression |  | 32 | 32 | 32 |  |  |  | no difference |
| Lindqvist et al. (2017) | Unipolar depression | Responders | 11 | 11 | NA | NA | 8 weeks | Psychotropics | no difference |
|  |  | Nonresponders | 8 | 8 | NA |  |  |  | increase |
| Tsai et al. (2016) | Unipolar depression |  | 18 | 18 | 9 | HDRS-17 scores≤7 | NA | Psychotropics | no difference |
| Abbreviation: HDRS, Hamilton Depression Rating Scale; MADRS, Montgomery Asberg Depression Rating Scale; NA, not available Bolt text indicates the studies included in the new dataset consisted of only remitted population data. | | | | | | |  |  |  |
